# Smart Investment of Virus RNA Testing Resources to Enhance Covid-19 Mitigation

**DOI:** 10.1101/2020.11.30.20239566

**Authors:** Hossein Gorji, Markus Arnoldini, David F. Jenny, Wolf-Dietrich Hardt, Patrick Jenny

## Abstract

Covid-19 mitigation commonly involves contact tracing (CT) and social distancing. Due to its high economic toll and its impact on personal freedom, we need to ease social distancing and deploy alternative measures, while preventing further waves of infections. While reliable mass testing (for virus RNA) would require too many resources to be effective, CT, which focuses on isolating symptomatic cases and their contacts, has been implemented in many countries. However, the latter approach has reduced efficiency when high numbers of positive patients are burdening the tracing centers. Moreover, CT misses transmissions by asymptomatic cases. Therefore, its effect in reducing the reproduction number has a theoretical limit.

To improve effectiveness of contact tracing, we propose to complement it with a strategy relying on identifying and testing symptom free subgroups with a significantly higher than average virus prevalence. We call this smart testing (ST). By testing everybody in these subgroups, in addition to symptomatic cases, also large fractions of pre- and asymptomatic persons can be identified, which enhances the effectiveness of contact tracing. High prevalence subgroups can be found in different ways, which are discussed in this paper. A particularly efficient way is via preselection using cheap and fast virus antigen tests, as proposed recently. Mathematical modeling quantifies the potential reduction of the reproduction number by such a two-stage ST strategy. In addition to global scenarios, also more realistic local applications of two-stage ST have been investigated, that is, within counties, institutions, schools, companies, etc., where members have internal as well as external contacts. All involved model parameters have been varied within realistic ranges and results are presented with probabilities. Even with the most pessimistic parameter set, these results suggest that the effect of two-stage ST on the reproduction number would clearly outweigh its economic cost. Two-stage ST is technically and logistically feasible. Further, it is locally effective also when only applied within small local subpopulations. Thereby, two-stage ST efficiently complements the portfolio of mitigation strategies, which allow easing social distancing without compromising public health.

**Single Sentence Summary:** Identification of high prevalence groups within subpopulations to enhance detection rate of Covid-19 infections by virus RNA tests combined with subsequent isolation.

## Introduction

The Covid-19 pandemic has evaded containment measures, both initially and after the first wave. Current responses have therefore shifted towards mitigating the effects. However, proven vaccines and therapies are still months to years away and the current capacity for detecting the virus via its genomic RNA is limited [1]. Thus, mitigation in many countries relies on a broad portfolio that includes not only hygiene measures, but also physical distancing (termed social distancing, hereafter) to reduce transmission, diagnosing virus (SARS-CoV2) infections in infected people showing mild to severe symptoms and tracing of their recent contacts. In combination, these measures slow down pandemic spread and help to avoid overburdening healthcare systems by easing the demand for intensive care. However, this strategy has two major shortcomings. First, it leaves many infected people with mild or no symptoms undetected [2], and therefore renders them more likely to infect others. Second, as social distancing measures limit non-essential business, it imposes severe economic consequences. A broad social-distancing-based approach is therefore not sustainable, but mitigation measures need to stay in place until effective therapies or vaccines become available to avoid further waves of virus spread. As these options are still months to years away, we need to consider alternative mitigation strategies.

Classical contact tracing and bluetooth-based smartphone apps can help to identify individuals that have recently had an infection-relevant contact (i.e., one that confers a risk of transmission) with known Covid-19 cases and might therefore have been infected. Self-quarantine of these contacts can help to mitigate the pandemic [3,4]. However, the effectiveness of contact tracing is limited by the participation, the efficiency in identifying infected people and relevant contacts, the delays in alerting contact persons and by important epidemiological characteristics, e.g. the fraction of asymptomatic cases and their infectiousness [3,4]. Infectious people who are not showing symptoms are limiting the efficiency of both, classical and app-based contact tracing: for example, the study of the COVID-19 outbreak in the municipality of Vo’, Italy conducted by Lavezzo et al. [5], performed two virus tests two weeks apart, and observed that 43.2% of infected people were asymptomatic and that their viral loads were as high as those of symptomatic cases. Contact tracing of symptomatic cases by bluetooth apps or traditional methods would have missed these cases and failed to detect and quarantine their contacts. Infectiousness of asymptomatic cases has been estimated to be between 10% and 100% of symptomatic ones [4-6] (based on differences in viral load), and the fraction of asymptomatic cases has been reported to be between 11.5% and 43.2% of all infected cases [6-8]. Asymptomatic cases are therefore a particular problem, as contact tracing detects them insufficiently and thereby limits the efficiency of this approach in breaking transmission chains and reducing the likelihood and impact of super-spreader events. In fact, we have recently shown analytically that contact tracing alone is insufficient to reduce R_eff_ to 1 even for the most optimistic assumptions [9]; contact tracing can therefore be a valuable mitigation tool for mitigation, but it needs to be complemented by other mitigation measures. We argue here, that this can be done efficiently, by focusing such efforts on detecting asymptomatic and presymptomatic infected people.

Recently, we have analyzed how mass testing random samples of the population and quarantine of positive cases could work to mitigate the pandemic. Importantly, by testing for virus RNA, a marker for active infections, also asymptomatic but infectious people can be detected. We estimated how many tests would be needed per day do mitigate the pandemic, i.e. to reduce the basic reproduction number R_0_ of the pandemic from 2.4-3 [10,11] to an effective reproduction number of R_eff_ =1. Unfortunately, however, the model shows that applying this approach non-selectively to the whole population at a reasonable frequency would require too many resources.

In this paper, we analyze an option to target the available numbers of virus RNA tests to high-prevalence subpopulations, a strategy we term smart testing (ST). These high-prevalence subpopulations are identified by prescreening with quick and cheap virus antigen tests, which detect structural components of the virus particles (not its RNA); This two-stage testing approach has recently been proposed [12], but has not yet been quantitatively analyzed. Our quantitative analyses indicate that such a two-stage ST strategy greatly reduces the required number of virus RNA tests and increases the mitigation power of testing in a tunable fashion.

## Model

To estimate how different mitigation strategies affect R_eff_, we use a mathematical model that employs a set of ordinary differential equations to describe the dynamics of the infections in a susceptible population. The model is an extension of the one used in [9]. The graph of the model is shown in **Fig. S1**, and it is described in detail in the SI, section 1. The model differs from standard epidemiological models in that individual compartments represent subpopulations before, during and after infection with respect to the detection of their current (or previous) infection. The gray field on the right represents those infected people that are already efficiently detected by current mitigation approaches. The insert on the top right shows a parallel graph of infected individuals which could be detected in addition, if virus testing is applied to subpopulations with no symptoms (or only mild symptoms). To obtain realistic simulation results, we parametrized our model using published data [6,13-18] (see SI section 2). Infection of susceptible people will lead to a latency phase. The exposed will later become transmissive, and either remain asymptomatic and recover or become pre-symptomatic and later develop mild symptoms. These two transmissive groups do not know that they are infected, and thus remain unidentified in current mitigation approaches. Some people with mild symptoms of disease will self-isolate. Due to self-isolation, they will have a reduced probability to infect others (we assume a 90% reduction). Infected people with severe symptoms are hospitalized, immediately isolated under strict quarantine and do not infect others. The same applies for anyone else who is tested virus-positive; which is indicated by the dashed arrows in **Fig. S1**. As our model consists of a set of ordinary differential equations, which become linear in the early stages of the pandemic (when nearly the whole population still is susceptible), we can now analytically test how particular mitigation strategies affect R_eff_. We would like to point out that the aim of this study is not to predict the course of the pandemic and associated uncertainties for certain countries or regions, but rather to serve as realistic proxy for possible scenarios, for which the effects of different mitigation strategies (and their combinations) can be investigated and demonstrated.

## Results and Discussion

### Testing-based mitigation strategies to replace social distancing

In order to compare different mitigation strategies to a baseline, we first re-capitulate what the model predicts, if we omit any mitigation, that is, for the worst-case scenario without any measures. We assume a value for R_0_ (the basic reproduction number in the unmitigated case) that is in the mid-range of published estimates, i.e., that one infected person infects 2.4 others in average (see **Fig. 1A**). Under these circumstances, 87% will either recover or die from the disease within ≈250 days, which compares well to the 81% predicted by Ferguson et al. [6] for UK and US populations in the absence of mitigation plans. If alternative (but plausible) values are assumed for R_0_, these numbers are going to change only by a factor 2 or less for the unmitigated case (data not shown). Thus, our basic predictions are robust, even if input parameters for the virus infection dynamics might be subject to change when more precise parameters became available. Importantly, we show that there always is a considerable population of infectious but undetected people (**Fig. 1A**, dashed blue curve), for who targeted mitigation measures such as contact tracing or self-quarantine will be ineffective, and who are thus more likely to transmit the disease.

**Figure 1.**
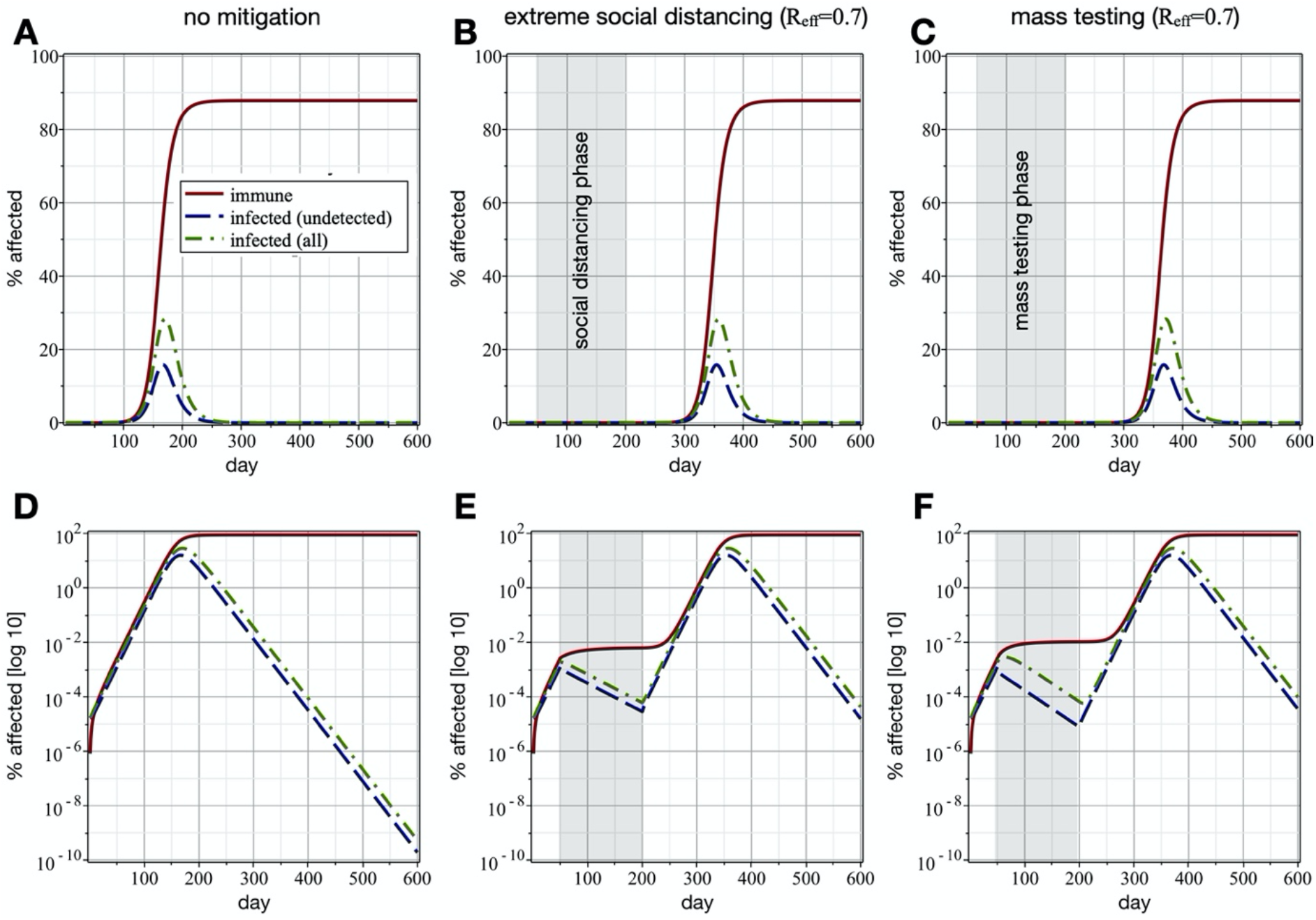
**Social distancing and extensive mass testing alone have qualitatively equal mitigating effects.** (A) Model outcome if no mitigation strategies are in place (R_0_=2.4). (B) Model outcome if extreme social distancing (71% lower infection rate, leading to R_eff_=0.7) is in place between days 50 and 200 of the outbreak. After day 200, social distancing is discontinued. (C) Model outcome if mass testing with isolation of detected cases is applied between days 50 and 200 of the outbreak (as shown in A). The shown effect is achieved if 37’400 people per 100’000 are tested every day (5% false negative rate; test speed = 1 day). Testing is discontinued after day 200. Solid red lines represent recovered plus infected plus deceased, dashed-dotted green lines infected, and dashed blue lines undetected infected people. Lower panels show the same data as upper panels, but with a log_10_-scale for Y-axes.

As the reproduction number scales linearly with the infection rate, reducing the infection rate by 71% via a generalized mitigation strategy such as social distancing leads to a decrease of the reproduction number by a factor of 3.4 (R_eff_ = 0.7). **Figure 1B** shows the course of the epidemic, if social distancing is imposed at this intensity for a period of 150 days (day 50-200 of the pandemic), whereas it is assumed that the basic reproduction number without any mitigation is R_0_=2.4. As expected, the number of infections decreases during the social distancing phase, but a new wave of infections starts as soon as mitigation is abandoned. These results are in line with current observations [3,4].

We have recently shown that testing random samples of the population with concomitant isolation of detected cases has a qualitatively identical effect as social distancing ([9] SMW manuscript, and SI section 3.1). For realistic assumptions of test processing times and quality of testing results, we found that 37’300 virus RNA tests per 100’000 people per day are required to reduce the reproduction number by a factor of 3.4, i.e., to achieve a similar effect on the epidemiological dynamic as reducing the infection rate by 71% via social distancing (compare **Figs. 1B** and **1C; Fig. S2** shows more generally how many tests are needed to achieve a given reduction factor in R_eff_, depending on test speed and quality). Deploying this number of tests every day is currently impossible for a variety of reasons, including availability of reagents, testing infrastructure, and compliance of the population. However, these results show that testing alone can mitigate the pandemic and that we can get a quantitative estimate for the number of tests needed in order to curb pandemic spread by testing alone.

### Mitigation via smart testing

Importantly, the number of tests needed to achieve the same result can be dramatically reduced, if the tests are not distributed randomly, but only subpopulations with higher infection prevalence are tested, and people who test positive are quarantined ([9], and SI section 3.2). We call such a strategy Smart Testing (ST).

For such a strategy to be successful in achieving R_eff_=1, the ratio of the subpopulation prevalence to the overall prevalence needs to be sufficiently high. In fact, the factor by which the number of required virus RNA tests can be reduced is equal to the ratio of prevalence in the test subpopulation to the overall prevalence [9]. **Figure 2** shows the number of required virus RNA tests per 100’000 people per day to achieve a specified reproduction number reduction factor for different prevalence ratios. For example, if the tested subpopulation has a 32 times higher prevalence than the overall population (a prevalence ratio of 32, then only 246 virus RNA tests per 100’000 people per day would be required to reduce the reproduction number by a factor of two (a 5% false negative rate and one day delay is assumed here for virus RNA tests). ST can be a realistic mitigation strategy, as the number of tests needed to achieve a sufficient reduction in R_eff_ is already available in several countries. The remaining key question, of course, is how to detect a subpopulation with a significantly increased prevalence. In general, such subpopulations can be identified based on correlations between infection probability and observable quantities. As these observables are often highly sensitive personal information, the type and amount of data needed to achieve the goal of identifying the high-prevalence subpopulation is crucial for it to be accepted by the citizens.

**Figure 2.**
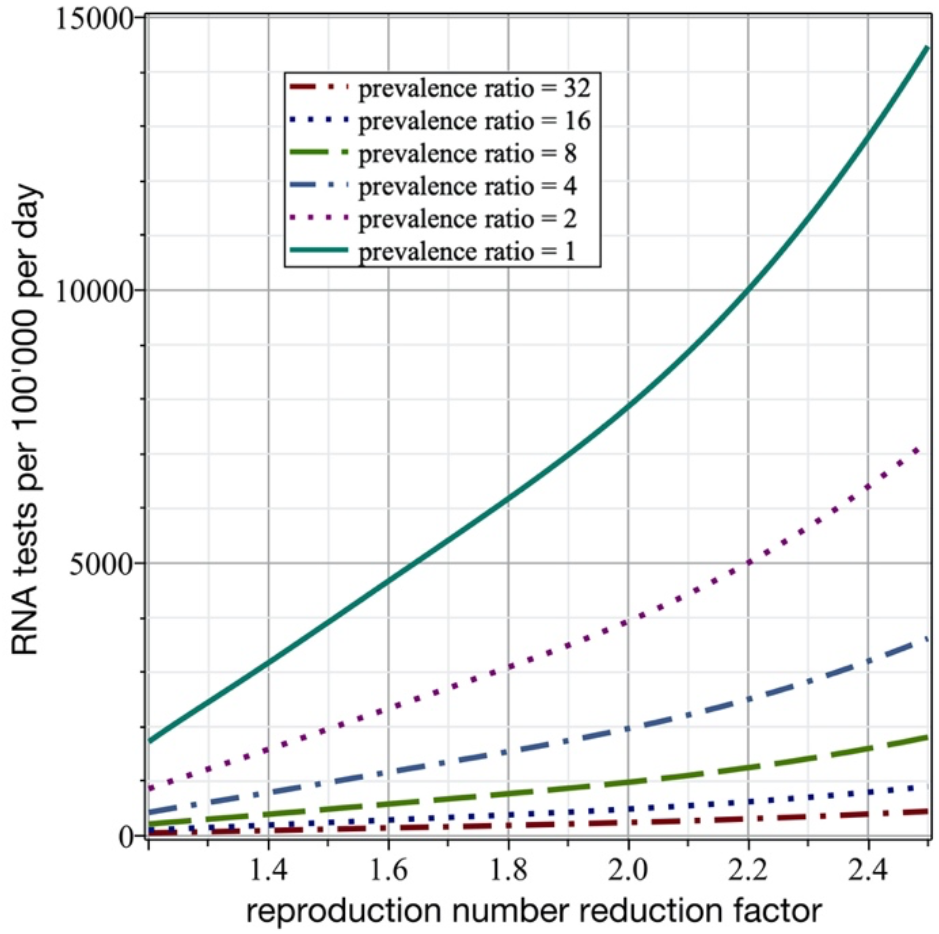
**Numbers of required virus RNA tests per 100’000 people per day for ST** to achieve a specified reproduction number reduction factor depending on the ratio of prevalence in the preselected test subpopulation and in the overall population. A 5% false negative rate and one day delay is assumed for the virus RNA tests.

### Identifying high-prevalence subpopulations

One possible strategy that could, in theory, be implemented in a privacy preserving way, relies on the fact that some individuals will have many more infection-relevant contacts than most others [19], and these high contact people are known to be highly important for epidemiological dynamics ([20], referred to as super-spreaders). Along these lines it was suggested to extend smartphone apps used for contact tracing by adding contact counting functionality for identifying high prevalence subpopulations. Mathematical modeling shows that smart testing with contact counting (STeCC) could work in realistic epidemiological scenarios [21], but the efficiency of such an approach is limited by the number of people using the app and by data privacy concerns.

Therefore, we investigate another way to identify high prevalence subpopulations. This approach is based on a preselection by virus antigen tests. Due to their relatively high false negative and false positive rates, these tests are controversial as stand-alone tests. However, if used as a pre-screening tool to identify high-prevalence subpopulations for subsequent virus RNA testing, they could be extremely useful despite their low specificity. In a strategy we call two-stage smart testing, random groups of people (i.e. those without disease symptoms or only mild symptoms at best) would be tested using the relatively cheap and easy to use virus antigen tests, and only people who are tested positive in this first stage would then be tested using a virus RNA test.

Depending on the false positive and false negative rates of the antigen test (f_p_ and f_n_, respectively), we can calculate the fraction of the population which gets preselected in the first round of tests as r_s_=p(1-f_n_)+(1-p)f_p_, where p is the prevalence in the undetected overall population. The factor by which the prevalence increases in the preselected subpopulation (the prevalence ratio) is then r_p_=(1-f_n_)/r_s_. For example, with the realistic values of f_n_=30% and f_p_=1%, and with p=0.3%, one obtains r_s_=0.012 and r_p_=58. Both values increase for lower false negative and false positive rates. Further, the prevalence ratio decreases for a higher p, which underlines the value of starting with this mitigation measure early in the pandemic. In the example considered here, merely 136 virus RNA tests per 100’000 people per day (a processing time of one day and a false negative rate of 5% for the virus RNA tests are assumed) would be required to reduce the reproduction number by a factor of two. Applying these 136 virus RNA tests to people from the preselected subpopulation has the same effect as virus RNA-testing 7’870 random persons without pre-selection. This dramatically smaller number of required virus RNA tests (r_p_=58 times fewer compared to blind mass testing) makes two-stage smart testing a promising mitigation strategy, if a high enough r_p_ can be achieved.

Should there be an antigen test with lower false positive and false negative rates, it could in theory be used as a stand-alone test for mass testing. However, developing tests that can detect low virus titers without an amplification step is expected to be challenging. If low-specificity antigen tests are used as a stand-alone solution to decide whether people should quarantine, this would lead to a large population of healthy people being forced to self-isolate (**Fig. S6**), potentially depressing overall compliance with mitigation measures. Therefore, tests with significant false-positive rates will in most cases have to be applied in two-stage testing schemes, as analyzed herein.

### Requirements for successful two-stage smart testing

The next obvious question is, how many virus antigen tests are required per day to achieve this?

We can estimate the most economical number of antigen tests by setting the condition that the thereby detected high-prevalence subpopulation has to be of exactly the right size to achieve the desired reduction in R_eff_: first, if r_ag_ is the fraction of the total population we need to pre-select for testing by antigen tests, and a fraction r_s_ of these will be tested positive and makes up the high prevalence subpopulation to be tested using RNA tests, the fraction of the total population that makes up the high prevalence subpopulation is r_ag_r_s_. Second, if r_mt_ is the fraction of the population that would need to be RNA tested in a random mass testing strategy to achieve the desired reduction in R_eff_ (based on our modeling results), and r_p_ is the factor by which prevalence increases in the high prevalence subpopulation (see above), the fraction of the population that needs to be tested in a two-stage smart testing strategy to achieve the same reduction in R_eff_ is r_mt_/r_p_. Third, we set the condition that the population fractions r_ag_r_s_ (the high prevalence subpopulation detected using antigen tests) and r_mt_/r_p_ (the population fraction that needed to achieve the desired reduction in R_eff_) need to be equal, and resolve for r_ag_. This leads to the simple expression r_ag_=r_mt_/(r_p_ r_s_)=r_mt_/(1-f_n_).

In summary, this elegant result implies that the fraction of the overall population to be tested with virus antigen tests every day is r_mt_/(1-f_n_) and the fraction of the overall population to which virus RNA tests have to be applied is r_mt_/r_p_=r_mt_ r_s_/(1-f_n_)=r_mt_(p+(1-p)f_p_/(1-f_n_)).

To give a concrete and realistic example, with an overall prevalence of 0.3%, this means that 11’240 antigen tests and 136 virus RNA tests are required per 100’000 people per day to reduce the reproduction number by a factor of two. If the overall prevalence is 0.1% (or 0.9%), then 120 (or 182) virus RNA tests per 100’000 people per day suffice to have the same effect on the reproduction number. The number of required antigen tests, on the other hand, is not affected by the overall prevalence. Assuming respective costs of 57.5CHF and 114.5CHF (current values for Switzerland [22], including all involved personnel charges) for each antigen and RNA test, one calculates that merely 6.6CHF have to be spent in average per person per day. This cost is extremely low considering the enormous gain in mitigation and the economic costs of alternative mitigation strategies. **Figures 3A** and **3D** show the numbers of required virus antigen and RNA tests per 100’000 people per day as functions of the reproduction number reduction factor and the overall prevalence p. Note that the number of required antigen tests is independent of the overall prevalence (**Fig. 3A**), while more virus RNA tests are required as p increases (**Fig. 3D**).

**Figure 3.**
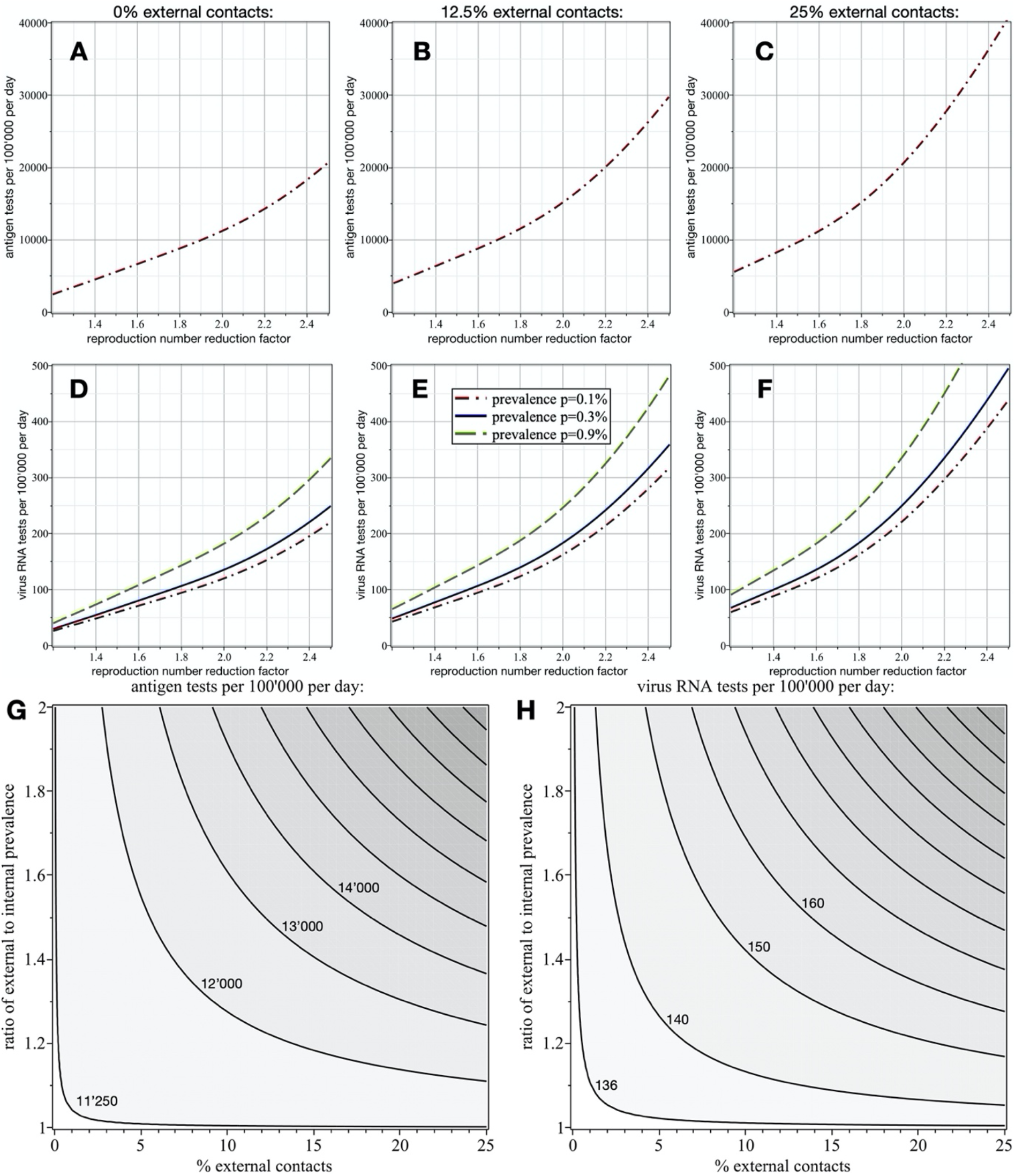
**Numbers of required tests** per 100’000 people per day as function of reproduction number reduction factor and prevalence p in the undetected population. In the first row (A-C) the number of antigen tests and in the second row (D-F) the number of virus RNA tests are shown. No external contacts are assumed for the results shown in the first column, while 12.5% and 25% external contacts are assumed for the plots in the second and third columns, respectively, where the external population has a two times higher prevalence. As expected, more tests are required to achieve the same reproduction number reduction as the fraction of external infection-relevant contacts increases. Also note that the number of required virus RNA tests increases with a higher overall prevalence (D-F), while the number of antigen tests is independent of p (A-C). Also shown are the effects of fraction of external contacts and ratio of external to internal prevalence on the numbers of required antigen (G) and RNA tests (H) per 100’000 people per day to reduce the reproduction number by a factor of two. A 5% false negative rate and one day delay is assumed for the virus RNA tests, and for the antigen tests false negative and false positive rates of 30% and 1% are assumed.

### Two-stage smart testing deployment strategies

We now have an estimate for the number of antigen and RNA tests needed to achieve a fast and strong reduction in R_eff_. But once the total number of cases in the population has declined sufficiently, testing can either be reduced or discontinued for a period of time before a new round of tests is initiated. In the following, we study the epidemiological consequences of different deployment strategies of two-stage ST.

**Figures 4A-C** show the overall prevalence and prevalence in the undetected population (dashed and solid lines, respectively), and **Figs. 4D-F** and **Figs. 4G-I** the number of deployed virus antigen and RNA tests per 100’000 people per day, respectively, as functions of time. The unmitigated reproduction number is 1.6 and each scenario starts on day 250, when the overall prevalence just exceeded 1%. The first scenario (first column) follows a two-stage ST strategy, in which for a first period of 50 days 18% of population is virus antigen tested every day. Once the prevalence is reduced by almost one order of magnitude, two-stage ST is continued at a lower intensity, that is, with 7% of the population being antigen tested every day. The second scenario (second column) is identical to the first one, except that the first phase lasts for 100 days, which leads to a reduction of the prevalence by almost two orders of magnitude. In the third scenario (third column), two-stage ST (with 18% of the population being antigen tested every day) is applied in cycles; each with 110 days of two-stage ST followed by a pause of 90 days. Monitoring could be done, for example, by using the relation p=(r_s_-f_p_)/(1-f_n_-f_p_), where r_s_ is the fraction of positive cases when virus antigen testing (with false negative and false positive rates of f_n_ and f_p_, respectively) is applied to a representative subpopulation.

**Figure 4.**
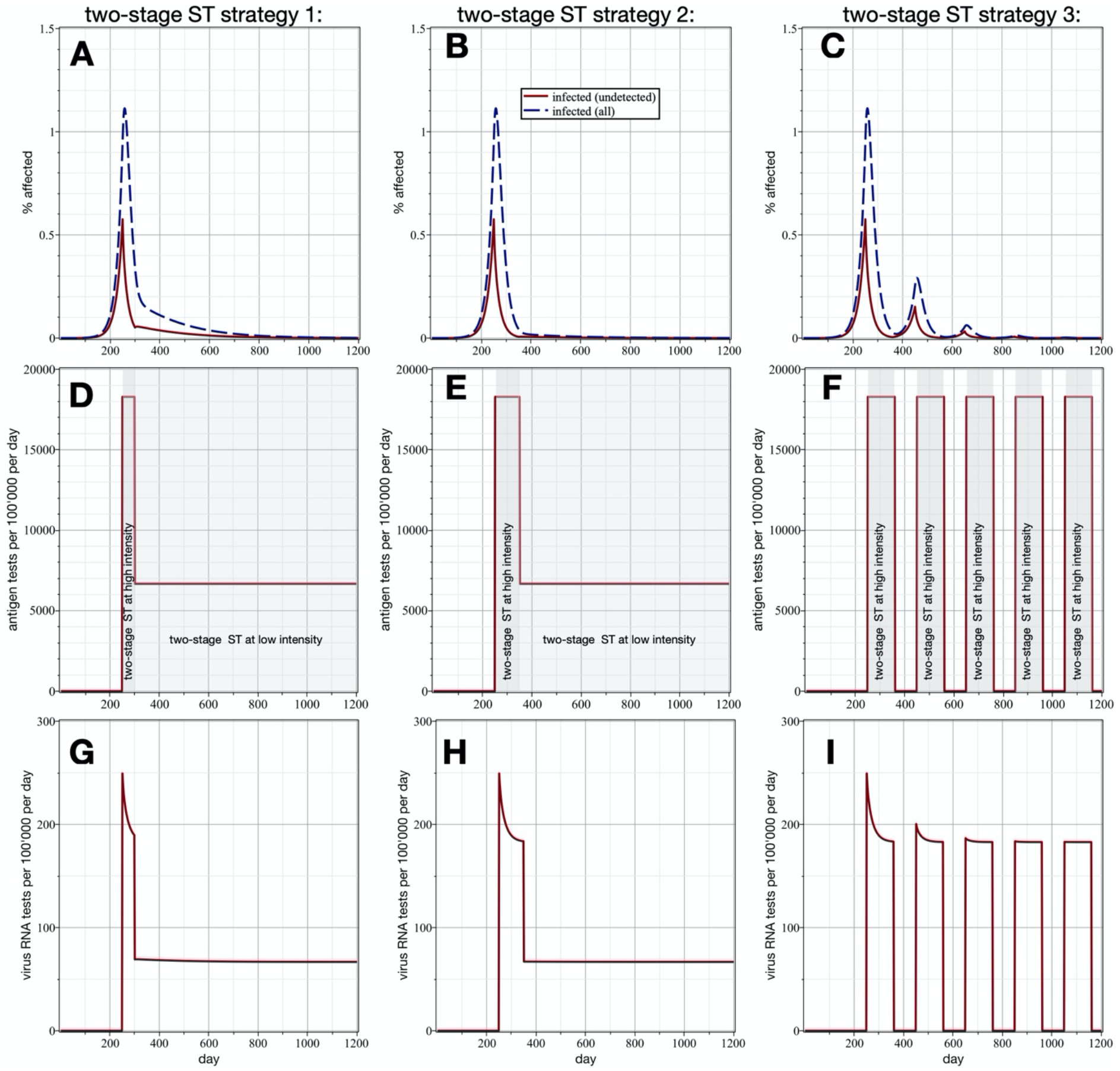
**Different two-stage ST mitigation strategies.** Shown are overall prevalence and prevalence in the undetected population (A-C; dashed and solid lines, respectively), and number of virus antigen and RNA tests per 100’000 people per day (D-F and G-I, respectively). The unmitigated reproduction number is 1.6 (already reduced by moderate social distancing) and each scenario starts on day 250, when the prevalence just exceeded 1%. The first scenario (first column) follows a two-stage ST strategy, in which for a first period of 50 days 18% of population is virus antigen tested every day. Once the prevalence is reduced by almost one order of magnitude, two-stage ST is continued at a lower intensity, that is, with 7% of the population being antigen tested every day. The second scenario (second column) is identical to the first one, except that the first phase lasts for 100 days, which leads to a reduction of the prevalence by almost two orders of magnitude. In the third scenario (third column), two-stage ST (with 18% of the population being antigen tested every day) is applied in cycles; each cycle starts with 110 days of two-stage ST followed by a 90 day pause. A 5% false negative rate and one day delay is assumed for the virus RNA tests, and for the antigen tests false negative and false positive rates of 30% and 1% are assumed.

Using the simple analysis above, it is straight forward to plan an effective two-stage ST campaign, and it is possible to predict its effect on R_eff_. The required numbers of antigen and virus RNA tests can directly be computed from known quantities (the overall prevalence is not known, but can be estimated based on positive antigen tests). To compensate for statistical noise and modeling uncertainties, we would advise, however, to choose slightly higher test numbers than the calculated ones. Naturally, the mitigation effect of two-stage ST can further be enhanced, if combined with other measures, such as contact tracing, mask wearing, or mild forms of social distancing.

So far, our analysis has focused on a closed population. Realistically, however, a two-stage ST strategy will always be employed on a subpopulation (e.g. in a country, state, city, school, or company), which is in constant exchange with other populations. Next, we therefore investigate scenarios in which two-stage ST is applied within subpopulations only, while no such measures are taken in the remaining population.

### Mitigation via two-stage smart testing within subpopulations

Generally, in a focal subpopulation in which a fraction r_ec_ of all infection relevant contacts happens with people external to that subpopulation, the virus reproduction number scales with the factor f_ec_=(1+r_ec_(r_ep_-1)), where r_ep_ is the ratio of external to internal prevalence. For example, if 25% of the infection relevant contacts are with an external population (r_ec_=0.25), which has a two times higher prevalence (r_ep_=2), then the reproduction number in the subpopulation is f_ec_=1.25 times higher than it would be with only internal contacts. This finding is very convenient to quantify the effects of two-stage ST, if restricted to a subpopulation. To obtain the numbers of required tests (and the resulting costs), one can simply use the results of two-stage ST in isolated populations (first column in Fig. 3) with the modified reproduction number (actual reproduction number without external contacts increased by the factor f_ec_). **Figs. 3B-C** and **Figs. 3E-F** show, for a two times higher external prevalence, the respective numbers of required virus antigen and RNA tests per 100’000 people per day as functions of the reproduction number reduction factor and the overall prevalence p for r_ec_=12.5% and r_ec_=25%. **Figure 3G** depicts the number of required antigen and **Fig. 3H** the number of RNA tests per 100’000 people per day to reduce the reproduction number by a factor of two, as functions of fraction of external contacts and ratio of external to internal prevalence. These results suggest that two-stage ST is a viable strategy to mitigate the pandemic, even on a relatively small scale.

## Conclusion

Two-stage ST adds to the portfolio of mitigation strategies for the Covid-19 pandemic, and complements approaches like classic contact tracing, hygiene measures, randomized testing of cohorts of interest, or other surveillance tools. It could be deployed quickly in countries with sufficient testing capacities like Switzerland (capacity ≈230 virus RNA tests per 100’000 per day). Importantly, we show that low specificity of the antigen tests is not inhibitory for the suggested strategy, opening up options that allow faster and cheaper deployment of such tests (e.g. testing by non-expert personnel, saliva instead of nose swabs). The earlier such a strategy is adopted, the less logistically and fiscally costly it will be. Once two-stage ST is implemented, one can adjust the strategy flexibly in order to ensure the desired performance. Two-stage ST offers a realistic approach to help relaxing broad social distancing policies in the near future without compromising health, while at the same time providing public health officials with much needed actionable information on the success of their interventions. This will be an important prerequisite for reclaiming our normal public life and initiating economic recovery.

## Materials and Methods

The dynamic model was implemented with Maple 2018. The calculations for mass testing, contact tracing and smart testing were implemented with MATLAB and the Statistics Toolbox Release 2018b. The corresponding codes are available on GitHub via https://github.com/gorjih2/STeCC_preliminary.

## Data Availability

All relevant data is included in the manuscript and supporting information.

## Acknowledgements

The authors are very thankful to Dario Ackermann, who created the website with the simulation tool based on the model presented in this paper. The authors would like to thank Emma Slack, Erik Bakkeren and Noemi Santamaria for helpful comments on the manuscript and Maxime Augier for constructive feedback on the technical implementation of STeCC. HG acknowledges funding from the Swiss National Science foundation (grant number 174060).

## Supplementary Materials

1. Supplementary Text describing the mathematical modeling approaches.

