## Supplementary material for "Smart Investment of Virus RNA Testing Resources to Enhance Covid-19 Mitigation"

### Supplementary Text for: Smart Investment of Virus RNA Testing Resources to Enhance Covid-19 Mitigation

#### 1. Model

Since pre-symptomatic and asymptomatic virus carriers play a central role in transmission of SARS-Cov-2 [8, 14, 19], it is necessary that the mathematical model accounts for both detected and undetected infected people. We developed a compartmental model as a generalized Susceptible-Exposed-Infectious-Recovered (SEIR) method [22, 12, 17, 10, 5], which besides honouring the mentioned distinction allows for studying cross-infections. The latter offers the opportunity to study mitigation scenarios localized to a sub-population e.g. cities, institutions, schools, etc. The basis of the model is similar to our previous work [9], where all disease relevant states are modeled as compartments. Below, first we briefly review the model and then investigate the cross-infections in more details. A corresponding graph is shown in Fig. S1. Consider a susceptible population $n_s$ at time $t$, with $n_s^0 = n_s(t = 0)$. Once the virus is contracted, the infected person is exposed and belongs to $n_e$. After the latency period, the exposed person becomes infectious. However depending on the way the virus triggers the immunity response, the person may remain asymptomatic or develop symptoms, whose populations are denoted by $n_{ia}$ and $n_{im}$, respectively. Once symptomatic persons feel the onset of symptoms, we assume that they seek self-isolation roughly after one day, and thus their infectiousness drops significantly. The population of symptomatic individuals in self-isolation is denoted by $n_{ms}$. Eventually, symptomatic persons recover or get hospitalized. We further make an approximation that hospitalized patients do not noticeably contribute to the disease propagation, and therefore their infectiousness is neglected. Consequently, the recovery or death of the hospitalized patients do not alter our analysis. This also makes our findings independent of the fatality rate, which is subject of dispute and has significant age stratification [3, 2, 18]. Table S1 provides a short description of the employed symbols.

The described mechanism can be cast into a set of ordinary differential equations, which governs the evolution of

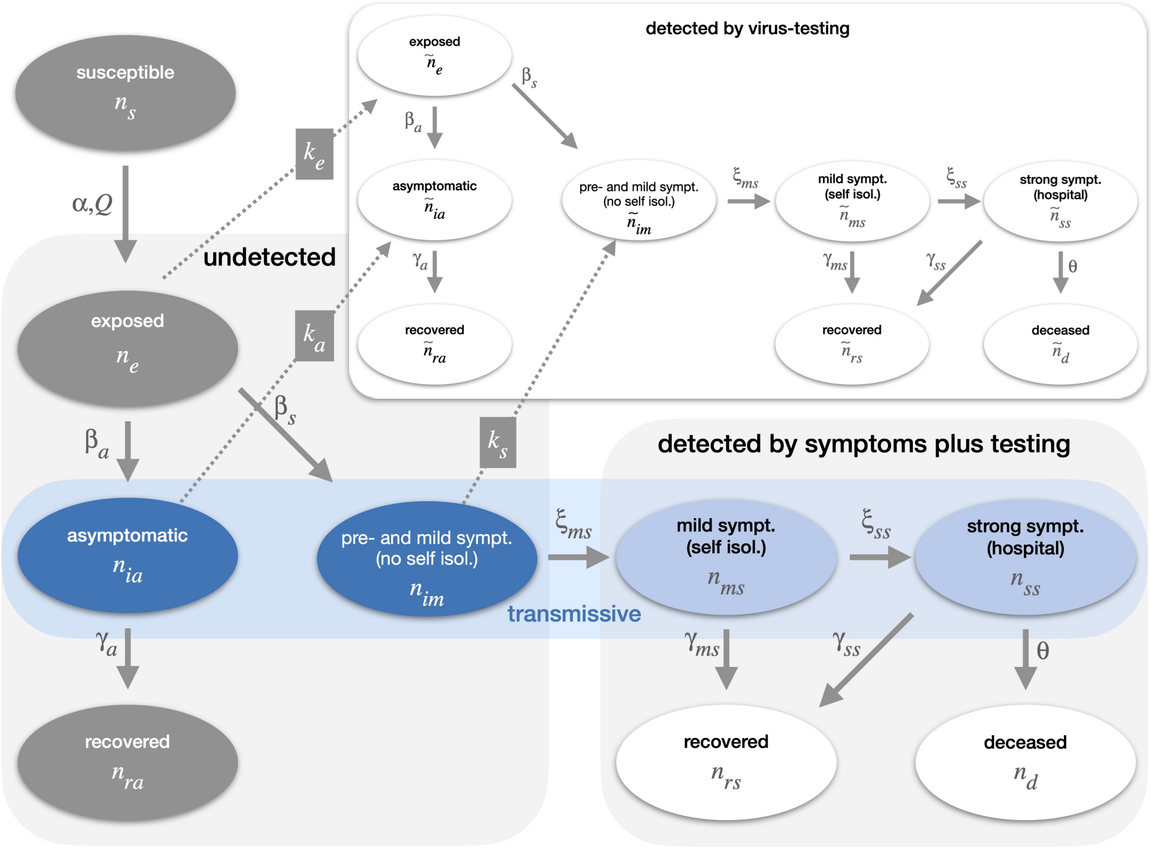


**Fig. S 1.** Graphical illustration of the modeling approach showing the dependencies within the system describing the dynamics of the susceptible, undetected and detected infected populations. It is crucial that the model distinguishes between individuals detected by symptoms (light blue), and those detected by virus testing (inserted graph). The detection rates of exposed, asymptomatic and mild symptomatic persons due to testing are proportional to $k_e$, $k_a$ and $k_s$, respectively. These individuals are then accounted for in the inserted graph with the white compartments, which is very similar as the main one, except that there is no node for susceptible persons (since by definition a susceptible person cannot be detected infected) and that there exist sources due to testing (dotted arrows) instead of sinks.

| **terminology** | **meaning** |
|---|---|
| susceptible | persons of the considered population who are susceptible and thus can potentially get infected |
| exposed | infected persons; can not yet transmit the virus |
| asymptomatic | infected persons without symptoms; can transmit the virus |
| pre- and mild sympt. (no self isol.) | infected persons with no or mild symptoms; infectious, but not isolated |
| mild sympt. (self isol.) | infected persons with mild symptoms; infectious and isolated |
| strong symptomatic | infected persons with strong symptoms and thus hospitalized; isolated |
| deceased | persons who died |
| recovered | persons who recovered |
| detected | isolated either after positive testing or after falling ill |
| undetected | persons who are either exposed, asymptomatic or mild symptomatic, but were never contained |
| transmissive | persons who are either asymptomatic or symptomatic |
| mild social distancing | $\mathcal{R}_{\text{eff}} = 1.6$, if the infection rate is reduced by 33% via social distancing |
| **variables** | |
| $n_s$ and $n_s^0$ | numbers of susceptible and initially susceptible persons, respectively |
| $n_e$, $\tilde{n}_e$ and $n_e^{tot}$ | numbers of exposed persons; not tested, tested and in total, respectively |
| $n_{ia}$, $\tilde{n}_{ia}$, $n_{ia}^{tot}$ and $n_{ia}^e$ | numbers of asymptomatic persons; not tested, tested, in total and in external population, respectively |
| $n_{im}$, $\tilde{n}_{im}$, $n_{im}^{tot}$ and $n_{im}^e$ | numbers of persons with mild symptoms during first day; not tested, tested, in total and in external population, respectively |
| $n_{ms}$, $\tilde{n}_{ms}$, $n_{ms}^{tot}$ and $n_{ms}^e$ | numbers of persons with mild symptoms after first day; not tested, tested, in total and in external population, respectively |
| $n_{ss}$, $\tilde{n}_{ss}$ and $n_{ss}^{tot}$ | numbers of persons with strong symptoms; not tested, tested and in total, respectively |
| $n_{ra}$, $\tilde{n}_{ra}$ and $n_{ra}^{tot}$ | numbers of recovered persons who had no symptoms; not tested, tested and in total, respectively |
| $n_{rs}$, $\tilde{n}_{rs}$ and $n_{rs}^{tot}$ | numbers of recovered or deceased persons who had symptoms; not tested, tested and in total, respectively |
| $n_i^{undet}$ | undetected infected persons: $n_{ie} + n_{ia} + n_{im}$ |
| $n_i^{det}$ | detected infected persons: $n_{ms} + n_{ss} + \tilde{n}_{ms} + \tilde{n}_{ss} + \tilde{n}_{ie} + \tilde{n}_{ia} + \tilde{n}_{im}$ |
| **parameters** | |
| $\alpha$ and $\tilde{\alpha}$ | rate coefficients for infection without and with cross-infections |
| $\epsilon$ | ratio between infection rate of self-isolated and non-quarantined symptomatic cases |
| $\beta_a$ and $\beta_s$ | rate coefficients for latency of asymptomatic and symptomatic cases, respectively |
| $\gamma_a$, $\gamma_{ms}$ and $\gamma_{ss}$ | rate coefficients for recovery |
| $\xi_{ms}$ and $\xi_{ss}$ | rate coefficients for successively stronger symptoms |
| $k_e$, $k_a$ and $k_s$ | rate coefficients accounting for testing |
| $r_{ec}$ | ratio of external to overall contacts |
| $p^e$ and $p$ | prevalence of external and main population, respectively |
| $p^h$, $p^l$ and $r_p$ | prevalence of high-prevalence sub-population, low-prevalence complement and their ratio, respectively |
| $\mathcal{R}_0$ | basic reproduction number without mitigation |
| $\mathcal{R}_{\text{eff}}$ | effective reproduction number with mitigation |
| $\mathcal{R}_{\text{eff}}^{wt}$ | effective reproduction number subject to testing |
| $N$ | testing interval |
| $\eta$ | fraction of virus-RNA false negative test results |
| $f_n$ and $f_r$ | fraction of antigen false negative and false positive test results, respectively |
| $r_{mt}$, $r_{rna}$ and $r_{ag}$ | fraction of population subject to mass-testing, RNA testing and antigen testing per day, respectively |
| $r_{s_1}$, $r_{s_2}$ and $r_s$ | fraction of antigen tested population with positive, negative and either results, respectively |
| **operators & functions** | |
| $\mathbb{E}(\cdot)$ | expectation |
| $\mathcal{P}_{\mathcal{A}}$ and Prob$\{\mathcal{A}\}$ | probability of the event $\mathcal{A}$ |
| $\sigma(\eta, \tau_{proc}, r_{mt})$ and $r_{wt}$ | reduction in the reproduction number due to the mass-testing without and with cross-infections, respectively |

**Table S 1.** Terminology and nomenclature of model parameters and variables each compartment population. Without accounting for infections via external contacts the system reads

$$\begin{aligned}
\dot{n}_s &= -\alpha(n_{ia}/2 + n_{im} + \epsilon n_{ms})\frac{n_s}{n_s^0} && (1)\\
\dot{n}_e &= \alpha(n_{ia}/2 + n_{im} + \epsilon n_{ms})\frac{n_s}{n_s^0} - (\beta_a + \beta_s)n_e - k_e n_e && (2)\\
\dot{n}_{ia} &= \beta_a n_e - \gamma_a n_{ia} - k_a n_{ia}, && (3)\\
\dot{n}_{im} &= \beta_s n_e - \xi_{ms} n_{im} - k_s n_{im}, && (4)\\
\dot{n}_{ra} &= \gamma_a n_{ia}, && (5)\\
\dot{n}_{ms} &= \xi_{ms} n_{im} - (\gamma_{ms} + \xi_{ss}) n_{ms}, && (6)\\
\dot{n}_{ss} &= \xi_{ss} n_{ms} - \gamma_{ss} n_{ss} \quad \text{and} && (7)\\
\dot{n}_{rs} &= \gamma_{ss} n_{ss} + \gamma_{ms} n_{ms}. && (8)
\end{aligned}$$

To account for the cross-infection between internal and external populations, a key parameter is the ratio of external to overall contacts $r_{ec}$ (see [6] for more detailed cross-infection models). Considering the susceptible and exposed compartments, their dynamics are then modified to

$$\begin{aligned}
\dot{n}_s &= -\alpha(1 - r_{ec})(n_{ia}/2 + n_{im} + \epsilon n_{ms})\frac{n_s}{n_s^0} - \alpha r_{ec}(n_{ia}^e/2 + n_{im}^e + \epsilon n_{ms}^e)\frac{n_s}{n_s^0} \quad \text{and} && (9)\\
\dot{n}_e &= \alpha(1 - r_{ec})(n_{ia}/2 + n_{im} + \epsilon n_{ms}) + \alpha r_{ec}(n_{ia}^e/2 + n_{im}^e + \epsilon n_{ms}^e) - (\beta_a + \beta_s)n_e - k_e n_e, && (10)
\end{aligned}$$

where the superscript $(\cdot)^e$ denotes populations in the corresponding external compartments. The above dynamics can become more compact once the concept of the prevalence ratio of external to internal populations, i.e., $p^e/p$, is employed. Note that here the prevalence is the fraction of the infectious population within undetected exposed, asymptomatic and pre/mild-symptomatic compartments. Therefore since we assume that the self-isolated and hospitalized patients play a negligible role in the dynamics of the pandemic, their contribution in the prevalence are neglected. Assuming that the prevalence ratio is homogeneous among infectious compartments, we get

$$\frac{n_{ia}^e}{n_{ia}} = \frac{n_{im}^e}{n_{im}} = \frac{n_{ms}^e}{n_{ms}} = \frac{p^e}{p}, \tag{11}$$

which leads to

$$\begin{aligned}
\dot{n}_s &= -\alpha\left(1 + r_{ec}\left(\frac{p^e}{p} - 1\right)\right)\left(n_{ia}/2 + n_{im} + \epsilon n_{ms}\right)\frac{n_s}{n_s^0} \quad \text{and} && (12)\\
\dot{n}_e &= \alpha\left(1 + r_{ec}\left(\frac{p^e}{p} - 1\right)\right)\left(n_{ia}/2 + n_{im} + \epsilon n_{ms}\right)\frac{n_s}{n_s^0} - (\beta_a + \beta_s)n_e - k_e n_e. && (13)
\end{aligned}$$

Now it becomes evident that the net effect of cross-infections can be seen as a modification

$$\tilde{\alpha} = \alpha\left(1 + r_{ec}\left(\frac{p^e}{p} - 1\right)\right) \tag{14}$$

of the infection rate $\alpha$.

Considering a mass-testing campaign, people from exposed, asymptomatic and pre/mild-symptomatic compartments are removed by the rates $k_e$, $k_a$ and $k_s$, respectively. The dynamics of the individuals identified through the testing then follows

$$
\begin{aligned}
\dot{\tilde{n}}_e &= -(\beta_a + \beta_s)\tilde{n}_e + k_e n_e, &(15)\\
\dot{\tilde{n}}_{ia} &= \beta_a \tilde{n}_e - \gamma_a \tilde{n}_{ia} + k_a n_{ia}, &(16)\\
\dot{\tilde{n}}_{im} &= \beta_s \tilde{n}_e - \xi_{ms} \tilde{n}_{im} + k_s n_{im}, &(17)\\
\dot{\tilde{n}}_{ra} &= \gamma_a \tilde{n}_{ia}, &(18)\\
\dot{\tilde{n}}_{ms} &= \xi_{ms} \tilde{n}_{im} - (\gamma_{ms} + \xi_{ss})\tilde{n}_{ms}, &(19)\\
\dot{\tilde{n}}_{ss} &= \xi_{ss} \tilde{n}_{ms} - \gamma_{ss} \tilde{n}_{ss} \quad \text{and} &(20)\\
\dot{\tilde{n}}_{rs} &= \gamma_{ss} \tilde{n}_{ss} + \gamma_{ms} \tilde{n}_{ms}. &(21)
\end{aligned}
$$

#### 2. Parameter Estimation

Our proposed model is closed once the rate coefficients are estimated. We take the values from reported data mainly from [7, 23] as summarized in Table 2 (see [9] for more details). Note that these values can easily be adapted, if more reliable data becomes available. It is to be emphasized that our aim is not to perform high-fidelity scenario predictions. Yet we are interested in using the model as a proxy to explore our mitigation scenarios, while at the same time taking the parameters from a realistic range.

Following [7], the latency time $x_l$ is estimated to be in average half a day shorter than the incubation. The latter is considered to be log-normally distribution with mean $5.84 \pm 2.98$ days [21, 16]. Furthermore $1/3$ of all infections are assumed to be asymptomatic [7]. In average we expect that in one day from onset of symptoms, the symptomatic individuals seek self-isolation. Next, the average onset-to-discharge time of clinical cases is estimated to be 22 days, leading to the average hospital treatment time of 11 days [23, 7]. For mild-symptomatic cases which are assumed to be 80% of symptomatic cases [15], this interval is considered to be half i.e. around 11 days from symptom onset to recovery which gives in total 11.5 days from end of the latency period. A similar recovery time of 11.5 days is estimated for asymptomatic cases. Different values are suggested for infectiousness of asymptomatic cases e.g. 2/3 in [7] and 1 in [13]. We consider asymptomatic cases to be 50% less infectious compared to symptomatic ones. The self-isolated patients are assumed to have 90% less infectiousness. The infectious rate is computed using the virus basic reproduction number $\mathcal{R}_0 = 2.6$ [7, 1].

As the mortality rate of hospitalized patients varies significantly based on the age group and health care system, we simply do not compute the death rate, and combine deceased and recovered ones into one category. Nevertheless as it becomes evident in the following, the deceased population would not have an effect in the dynamics of the pandemic. To compute the virus basic reproduction number $\mathcal{R}_0$, we split the dynamics of the infected population into the infection driven propagation $f$ and the remainder $V$, i.e.,

$$
\begin{bmatrix} \dot{n}_e \\ \dot{n}_{ia} \\ \dot{n}_{im} \\ \dot{n}_{ms} \\ \dot{n}_{ss} \end{bmatrix} = \overbrace{\begin{bmatrix} 0 & \tilde{\alpha}/2 & \tilde{\alpha} & \epsilon\tilde{\alpha} & 0 \\ 0 & 0 & 0 & 0 & 0 \\ 0 & 0 & 0 & 0 & 0 \\ 0 & 0 & 0 & 0 & 0 \\ 0 & 0 & 0 & 0 & 0 \end{bmatrix}}^{f} \begin{bmatrix} n_e \\ n_{ia} \\ n_{im} \\ n_{ms} \\ n_{ss} \end{bmatrix} - \overbrace{\begin{bmatrix} \beta_a + \beta_s & 0 & 0 & 0 & 0 \\ -\beta_a & \gamma_a & 0 & 0 & 0 \\ -\beta_s & 0 & \xi_{ms} & 0 & 0 \\ 0 & 0 & -\xi_{ms} & \xi_{ss} + \gamma_{ms} & 0 \\ 0 & 0 & 0 & -\xi_{ss} & \theta + \gamma_{ss} \end{bmatrix}}^{V} \begin{bmatrix} n_e \\ n_{ia} \\ n_{im} \\ n_{ms} \\ n_{ss} \end{bmatrix}. \qquad (22)
$$

Note that testing is not considered here. The $\mathcal{R}_0$ of this system is the spectral radius of $fV^{-1}$, i.e.

$$\mathcal{R}_0 \;=\; \rho(fV^{-1}) \;=\; \frac{\tilde{\alpha}\beta_s}{\beta_a+\beta_s}\left[\frac{\beta_a}{2\gamma_a\beta_s}+\frac{1}{\xi_{ms}}+\frac{\epsilon}{\gamma_{ms}+\xi_{ss}}\right]. \tag{23}$$

For the scenario $\mathcal{R}_0 \leq 1$, the pandemic reaches an endemic state. From Eq. (23) we can see that the combined recovery and death rate of the hospitalized patients $\gamma_{ss}$ do not contribute to the reproduction number. This is the case as the virus transmission from hospitalized cases are assumed to be negligible.

Non-pharmaceutical interventions try to achieve an effective reproduction number $\mathcal{R}_{\text{eff}}$ which is significantly lower than $\mathcal{R}_0$. The most common approach so far has been social-distancing aiming at reducing $\alpha$ (and thus $\tilde{\alpha}$) by decreasing the average number of contacts. For mild social-distancing scenarios it is reasonable to assume that a reproduction number of 1.6 can be achieved. In the following we show how mass-testing and two-stage smart testing can further reduce the reproduction number below 1 and thus contain the virus spread.

| **parameters** | **value** |
|---|---|
| $\alpha$ | 0.670 (1/day) |
| $\epsilon$ | 0.1 |
| $\beta_a$ | 0.078 (1/day) |
| $\beta_s$ | 0.156 (1/day) |
| $\gamma_a$ | 0.087 (1/day) |
| $\xi_{ms}$ | 0.667 (1/day) |
| $\gamma_{ms}$ | 0.08 (1/day) |
| $\xi_{ss}$ | 0.02 (1/day) |
| $\gamma_{ss}$ | 0.091 (1/day) |
| **initial condition** | **value** |
| $n_e(0)/n_s^0$ | 1.5663e-07 |

**Table S 2.** List of estimated parameters and initial values. Note that our model allows to easily replace any of these parameters by more precise estimates, as more data become available. The initial values of all numbers except $n_e$ are set to zero.

#### 3. Mitigation via Testing

Motivated by the fact that asymptomatic and pre-symptomatic infected people significantly contribute to the propagation of the pandemic [8, 14], testing can provide a versatile mitigation approach. The idea is to invite a significant fraction of the susceptible population to mass-testing, and then ask the positively tested individuals to go into isolation. In order to quantify the consequence of this mitigation strategy on the effective reproduction number, one has to first compute the detection rates $k_e$, $k_a$, $k_s$. The main parameters that affect these rates are the number of conducted tests per unit of time, the fraction of false negatives and the processing time which includes both processing the results and notice-to-quarantine time delay. We neglect further virus transmissions from individuals isolated due to the test result. Below we first provide a general framework for computing these rates. Then we quantify a mass-testing scenario and finally integrate the idea of two-stage testing into our analysis. The first part has already been introduced in [9], and thus only the main results are presented here.

##### 3.1. Mass Testing

Suppose each susceptible person is scheduled to be tested once every $N$ days. Let $\tau_{\text{proc}}$ be the processing time, $\eta$ the fraction of false negatives and $\tau_{\text{det}}$ the detection time. The latter is assumed to be uniformly distributed in the interval $[1, N]$. Therefore the time delay between the infection at time $x \in [1, N]$ and detection would be $\tau_{det} = N - x + \tau_{proc}$. Furthermore, the latency time $x_l$ is taken to be log-normally distributed $5.34 \pm 2.7249$ (see Sec. 2). By comparing detection and latency times, the infected individual can be detected from one of the following compartments:

1. *Exposed:* Consider the event $\mathcal{A}$ when detection occurs during the latency period. Therefore the rate of detection from the exposed compartment becomes

$$k_e = (1-\eta)\mathcal{P}_{\mathcal{A}}\mathbb{E}_{\mathcal{A}}\left[\tau_{det}^{-1}\right]. \tag{24}$$

2. *Pre- and Mild-Symptomatic:* As people in this compartment eventually go into self-isolation after an average of one and half days since onset of symptoms, the relevant event set here is $\mathcal{B} : (\tau_{det} \geq x_l) \cap (\tau_{det} \leq (x_l + 3/2))$, from which one obtains

$$k_s = \frac{2}{3}(1-\eta)\mathcal{P}_{\mathcal{B}}\mathbb{E}_{\mathcal{B}}\left[\tau_{det}^{-1}\right]. \tag{25}$$

   Note that the factor $2/3$ accounts for the assumption that $2/3$ of infected individuals eventually develop symptoms.

3. *Asymptomatic:* Besides set $\mathcal{B}$, also those who remain asymptomatic, i.e., set $C : ((\tau_{det} \geq (x_l + 3/2)) \cap (\tau_{det} \leq (x_l + 11.5))$, are relevant, where a recovery time of 11.5 days is considered (see Sec. 2). Therefore the rate of detection from the asymptomatic compartment can be described as

$$k_a = \frac{1}{3}(1-\eta)\mathcal{P}_{\mathcal{B}}\mathbb{E}_{\mathcal{B}}\left[\tau_{det}^{-1}\right] + (1-\eta)\mathcal{P}_{C}\mathbb{E}_{C}\left[\tau_{det}^{-1}\right]. \tag{26}$$

Following an approach similar to that of Sec. 2, the effective reproduction number can be computed as

$$\mathcal{R}_{\text{eff}}^{wt} = \frac{\tilde{\alpha}\beta_s}{\beta_a + \beta_s + k_e}\left[\frac{\beta_a}{2(\gamma_a + k_a)\beta_s} + \frac{1}{\xi_{ms} + k_s}\left(1 + \frac{\epsilon\xi_{ms}}{\gamma_{ms} + \xi_{ss}}\right)\right]. \tag{27}$$

Based on Eq. (27) we can estimate the number of required tests per day for a given reduction in the reproduction number (reproduction number reduction factor), i.e., $\mathcal{R}_{\text{eff}}^{wt}/\mathcal{R}_0$, as a function of test characteristics and cross-infections. We consider a RNA mass-testing scenario for different test processing times, i.e., for $\tau_{proc} \in \{0.5, 1, 1.5\}$ days, and different false negative rates, i.e., for $\eta \in \{0.05, 0.15\}$. While it is observed that sensitivity and specificity of RNA tests depend on the time passed from infection [4], here fore simplicity we consider a constant fraction of false negative results among all tested compartments. Furthermore the cross infections are characterized by the fraction of external contacts $r_{ec} \in \{0\%, 12.5\%, 25\%\}$, where the prevalence ratio of external vs. internal population is assumed to be $p^e/p = 2$. We applied a Monte-Carlo technique to compute the detection rates. The results for the given parameter set are shown in Fig. S2 for the case without cross infections, and in Fig. S3(a) and (b) for the ratio of external contacts 12.5% and 25%, respectively. We observe from Fig. S2 that even in the absence of cross-infection, i.e., if mass-testing is applied to the whole population, the number of required tests exceed current capacities. While mass testing of $> 500 - 1'000$ per 100'000 people per day could be realized by taking advantage of next-generation RNA extraction combined with reverse transcription and PCR (see e.g. [11]), current virus RNA testing capacities in continental Europe hardly exceed 300 tests per 100'000 people per day. Therefore the required number of tests obtained by our estimations is still one order of magnitude larger than existing capabilities.

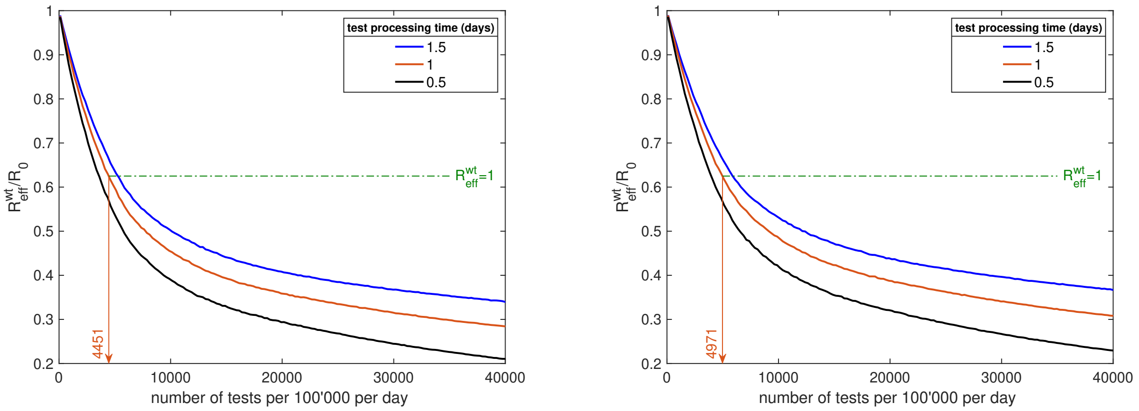


(a) (b)

**Fig. S 2.** Mass-testing: A mitigation strategy relying on RNA mass-testing is assumed and we computed the number of tests performed per day, which are needed to achieve a particular test-speed dependent $\mathcal{R}_{\text{eff}}^{wt}/\mathcal{R}_0$ ratio; for (a) 5% and for (b) 15% false negative test results are assumed. Test speeds were: 0.5 days (black line), 1 day (orange line) and 1.5 days (blue line). The green dashed line indicates reduction of reproduction number from 1.6 to $\mathcal{R}_{\text{eff}}^{wt} = 1$ via mass-testing.

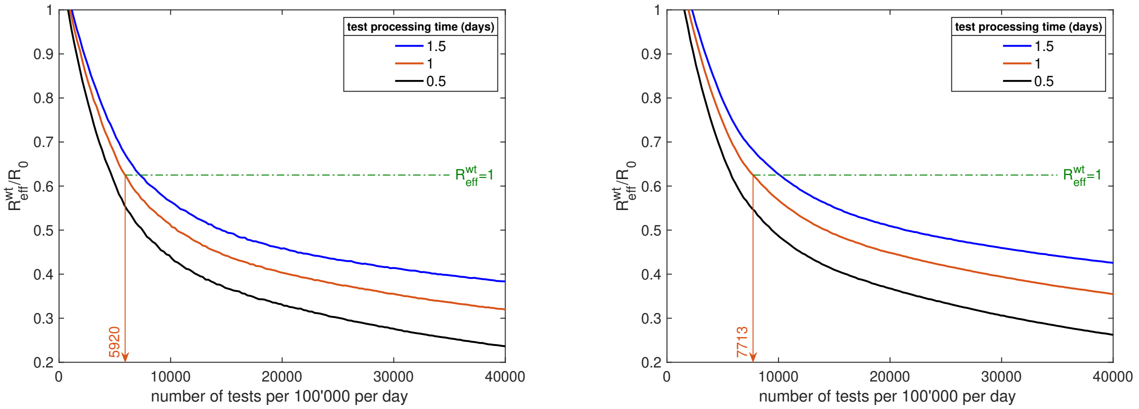


(a) (b)

**Fig. S 3.** Mass-testing: A mitigation strategy relying on RNA mass-testing alone is assumed and we computed the number of tests performed per day, which are needed to achieve a particular test-speed dependent $\mathcal{R}_{\text{eff}}^{wt}/\mathcal{R}_0$ ratio at 5% false negative test results; for (a) 12.5% and for (b) 25% external contacts. The prevalence ratio of 2 between the external and internal populations is considered. Test speeds were: 0.5 days (black line), 1 day (orange line) and 1.5 days (blue line). The green dashed line indicates reduction of reproduction number from 1.6 to $\mathcal{R}_{\text{eff}}^{wt} = 1$ via mass-testing.

Once we find the detection rates for a given test processing time and ratio of false negatives, computations for different scenarios can be carried out in a straight-forward way. Therefore it is more convenient to express the obtained results via a polynomial fit. More specifically, the reduction factor in the reproduction number can be written as

$$r_{wt} \quad = \quad \frac{\mathcal{R}_{\text{eff}}^{wt}}{\mathcal{R}_0} = \left(1 + r_{ec}\left(\frac{p^e}{p} - 1\right)\right)\sigma(\tau_{proc}, \eta, r_{mt}), \tag{28}$$

where $r_{mt}$ is the fraction of the susceptible population invited to be tested per day. An eighth order polynomial fit

$$\sigma \quad = \quad \sum_{i=0}^{8} a_i(\tau_{\text{proc}}, \eta)\, r_{mt}^i \tag{29}$$

is applied as the coefficients are tabulated in Table. 3.

| $\tau_{\text{proc}}$ [days] | $\eta$ | $a_0$ | $a_1$ | $a_2$ | $a_3$ | $a_4$ | $a_5$ | $a_6$ | $a_7$ | $a_8$ |
|---|---|---|---|---|---|---|---|---|---|---|
| 1 | 0.05 | 0.9900 | -11.5533 | 95.7615 | -448.4235 | 1238.5 | -2060.5 | 2026.9 | -1084.5 | 243.0903 |
| 1 | 0.15 | 0.9951 | -10.8494 | 88.5242 | -411.4751 | 1130.6 | -1873.4 | 1836.6 | -979.9444 | 219.1166 |
| 0.5 | 0.05 | 0.9808 | -13.2246 | 114.5028 | -552.5025 | 1557.6 | -2629.8 | 2615.1 | -1410.9 | 318.2973 |
| 0.5 | 0.15 | 0.9875 | -12.4584 | 105.7445 | -505.3994 | 1415.9 | -2379.3 | 2357.2 | -1267.9 | 285.3088 |
| 1.5 | 0.05 | 0.9942 | -10.3715 | 83.4762 | -383.6843 | 1048 | -1732.2 | 1697.2 | -905.9138 | 202.7611 |
| 1.5 | 0.15 | 0.9976 | -9.6937 | 76.8622 | -350.2807 | 950.2767 | -1561.4 | 1522.1 | -808.8726 | 180.3481 |

**Table S 3.** List of eighth order polynomial coefficients fitted to mass-testing results

##### *3.2. Two-Stage Smart Testing*

The efficiency of mass-testing can be significantly increased if a sub-population with a prevalence higher than in the total population can be found. In particular, consider a sub-population with prevalence $p^h$, whereas $p^l$ is the overall prevalence. It is straight-forward to show that the number of tests required to achieve the same effective reproduction number reduces by the factor

$$r_p \quad = \quad \frac{p^h}{p^l}; \tag{30}$$

see [9] for details. One approach to identify a high-prevalence sub-population is to deploy rapid antigen tests [20]. This of course comes with the price of large false negative and false positive results. Let us quantify how such a pre-selection can lead to a relevant mitigation scenario. Consider a population with a prevalence $p^l$, to which one applies antigen mass-testing with false negative and false positive ratios of $f_n$ and $f_p$, respectively. The select sub-population is composed of a fraction

$$r_{s_1} \quad = \quad (1 - f_n)p^l \tag{31}$$

of positive cases and a fraction

$$r_{s_2} \quad = \quad f_p(1 - p^l) \tag{32}$$

of negative cases. Therefore, the fraction of the screened sub-population is

$$r_s \quad = \quad r_{s_1} + r_{s_2} = p^l(1 - f_n) + f_p(1 - p^l), \tag{33}$$

and its prevalence is

$$p^h \quad = \quad \frac{(1 - f_n)p^l}{r_s}, \tag{34}$$

leading to a prevalence ratio of

$$r_p \quad = \quad \frac{1 - f_n}{r_s}. \tag{35}$$

Now, if we apply the virus RNA mass-testing strategy on the pre-screened sub-population, we achieve the same mitigation impact as without pre-screening, but with $r_p$ times fewer RNA tests. Consider a scenario where we invite a random fraction $r_{ag}$ of the population per day for antigen testing. A fraction $r_s$ of this population will be pre-screened based on the positive antigen test results. Therefore, compared to the original population, the fraction of the pre-screened sub-population is $r_{ag}r_s$. This fraction should match the number of required RNA tests relative to the overall population, i.e., $r_{mt}/r_p$, leading to the fraction

$$r_{ag} = \frac{r_{mt}}{1 - f_n} \tag{36}$$

of the susceptible population to be daily tested by antigen tests, and the fraction

$$r_{rna} = \frac{r_{mt}}{r_p} = r_{mt}\left(p^l + \frac{(1 - p^l)f_p}{1 - f_n}\right) \tag{37}$$

of the susceptible population to be daily tested by RNA tests.

To illustrate the performance of the two-stage testing mitigation, let us consider a concrete example. Suppose that the available antigen tests have false negative and false positive rates of $f_n = 30\%$ and $f_p = 1\%$, respectively. Furthermore, suppose that the pandemic is at an early stage with $p^l = 0.3\%$. The RNA tests are assumed to have false negative ratio of 5% and the total processing time is considered is $\tau_{\text{proc}} \in \{0.5, 1, 1.5\}$ days. We also consider cross-infections via contact with an external population, which has a two times higher prevalence, where the ratio of external to overall contacts is $r_{ex} \in \{0, 0, 12.5, 25\}$. Figures S4 (a) & (b) show results for the required numbers of antigen and RNA tests, respectively, for the case without cross-infection. The number of required RNA tests is reduced by the factor 58 compared to the simple mass-testing strategy. Furthermore, Figs. S5 (a) & (b) show the respective numbers of required antigen and RNA tests for 12.5% external contacts, and Figs. S4 (c) & (d) show the same data for 25% external contacts. As expected, more mixing with higher prevalence external population puts more burden on the numbers of tests. Nevertheless, the numbers of required antigen and RNA tests still are in the range of available resources. Figure S6 shows the fraction of infected cases among all people with positive virus antigen test result as function of prevalence; the left plot for 15%, 30% and 45% false negative rates and a fixed false positive rate of 1%; the right plot for 0.5%, 1% and 1.5% false positive rates and a fixed false negative rate of 30%. The fraction of actually infected cases among positively tested individuals can be calculated as $p(1 - f_n)/(p(1 - f_n) + (1 - p)f_p)$.

#### 4. Model Implementation

The calculations for mass-testing and two-stage testing were implemented with MATLAB and the Statistics Toolbox Release 2018b. The corresponding codes are available on GitHub server via `https://github.com/gorjih2/STeCC_preliminary`.

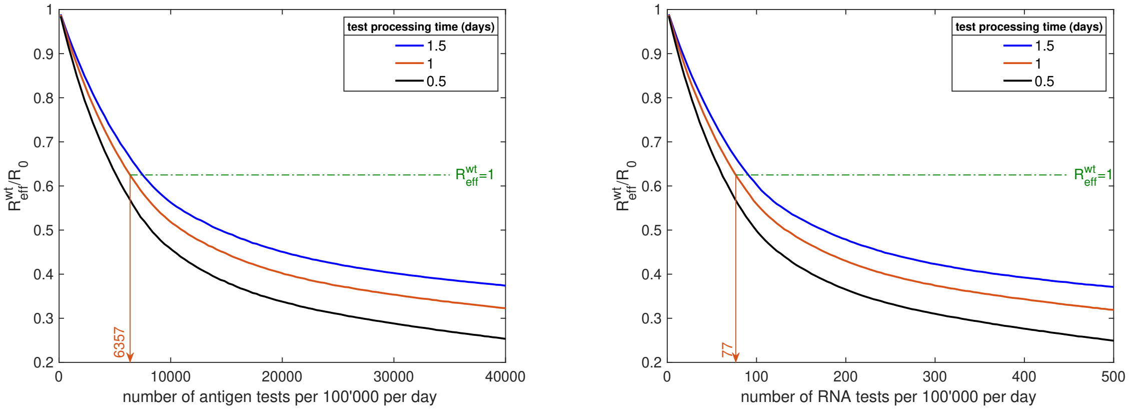


(a) (b)

**Fig. S 4.** Two-stage testing: A mitigation strategy relying on pre-screening using mass antigen testing and then RNA testing on the positive cases is assumed, and we computed the number of tests performed per day, which are needed to achieve a particular test-speed dependent $\mathcal{R}^{wt}_{\text{eff}}/\mathcal{R}_0$ ratio at 5% false negative RNA test results, 30% and 1% false negative and false positive antigen test results, respectively, and an overall prevalence of 0.3%. Combined test-to-quarantine speeds are 0.5 days (black line), 1 day (orange line) and 1.5 days (blue line).

[2] Andrew Atkeson. How deadly is covid-19? understanding the difficulties with estimation of its fatality rate. Technical report, National Bureau of Economic Research, 2020.

[3] Manuel Battegay, Richard Kuehl, Sarah Tschudin-Sutter, Hans H Hirsch, Andreas F Widmer, and Richard A Neher. 2019-novel coronavirus (2019-ncov): estimating the case fatality rate–a word of caution. *Swiss medical weekly*, 150(0506), 2020.

[4] Beatriz Böger, Mariana M Fachi, Raquel O Vilhena, Alexandre de Fátima Cobre, Fernanda S Tonin, and Roberto Pontarolo. Systematic review with meta-analysis of the accuracy of diagnostic tests for covid-19. *American journal of infection control*, 2020.

[5] Fred Brauer. Compartmental models in epidemiology. In *Mathematical epidemiology*, pages 19–79. Springer, 2008.

[6] Emmanuel J Dansu and Hiromi Seno. A model for epidemic dynamics in a community with visitor subpopulation. *Journal of theoretical biology*, 478:115–127, 2019.

[7] Neil M Ferguson, Daniel Laydon, Gemma Nedjati-Gilani, Natsuko Imai, Kylie Ainslie, Marc Baguelin, Sangeeta Bhatia, Adhiratha Boonyasiri, Zulma Cucunubá, Gina Cuomo-Dannenburg, et al. Impact of non-pharmaceutical interventions (npis) to reduce covid-19 mortality and healthcare demand. *London: Imperial College COVID-19 Response Team, March*, 16, 2020.

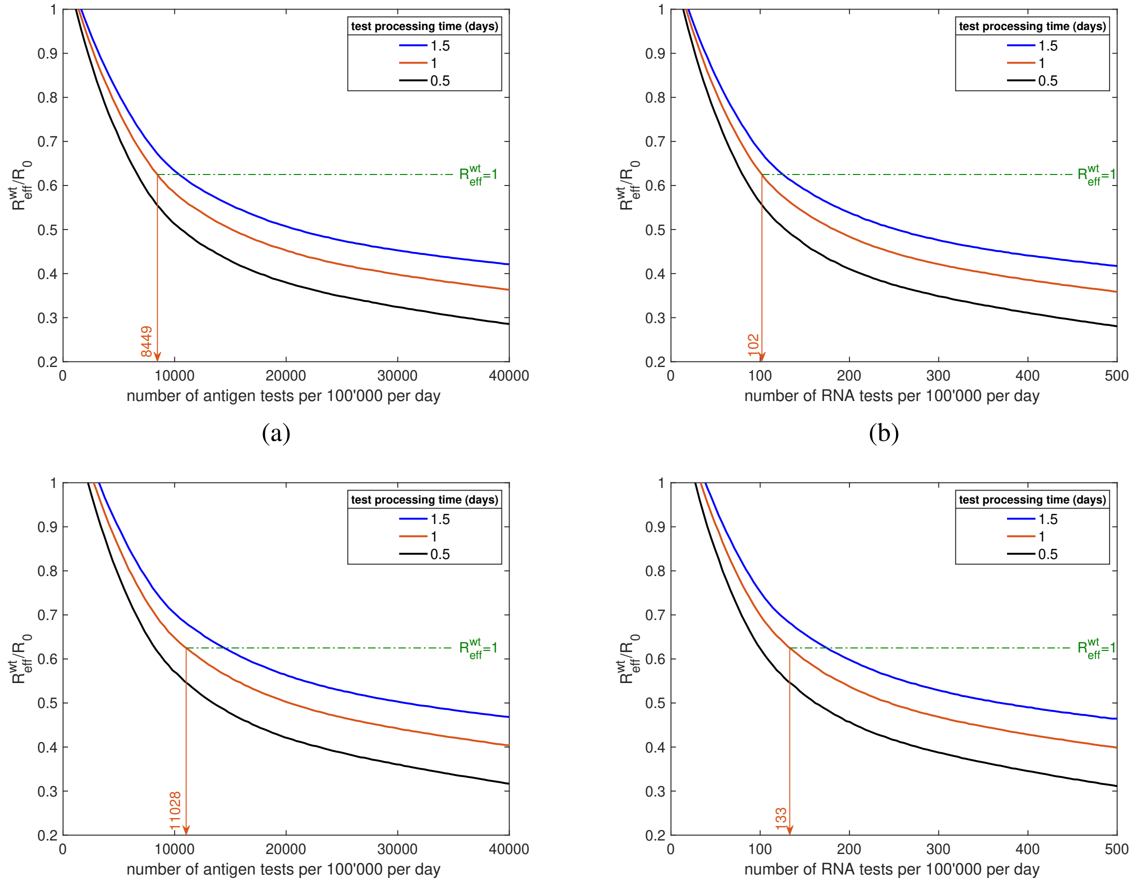


(a) (b) (c) (d)

**Fig. S 5.** Two-stage testing: A mitigation strategy relying on pre-screening using mass antigen testing and then RNA testing on the positive cases is assumed, and we computed the number of tests performed per day, which are needed to achieve a particular test-speed dependent $\mathcal{R}_{\text{eff}}^{wt}/\mathcal{R}_0$ ratio at 5% false negative RNA test results, 30% and 1% false negative and false positive antigen test results, respectively, and overall an prevalence of 0.3%; for (a) and (b) with 12.5% external contacts; for (c) and (d) with 25% external contacts. A prevalence ratio of 2 between the external and internal populations is assumed. Combined test-to-quarantine speeds were 0.5 days (black line), 1 day (orange line) and 1.5 days (blue line).

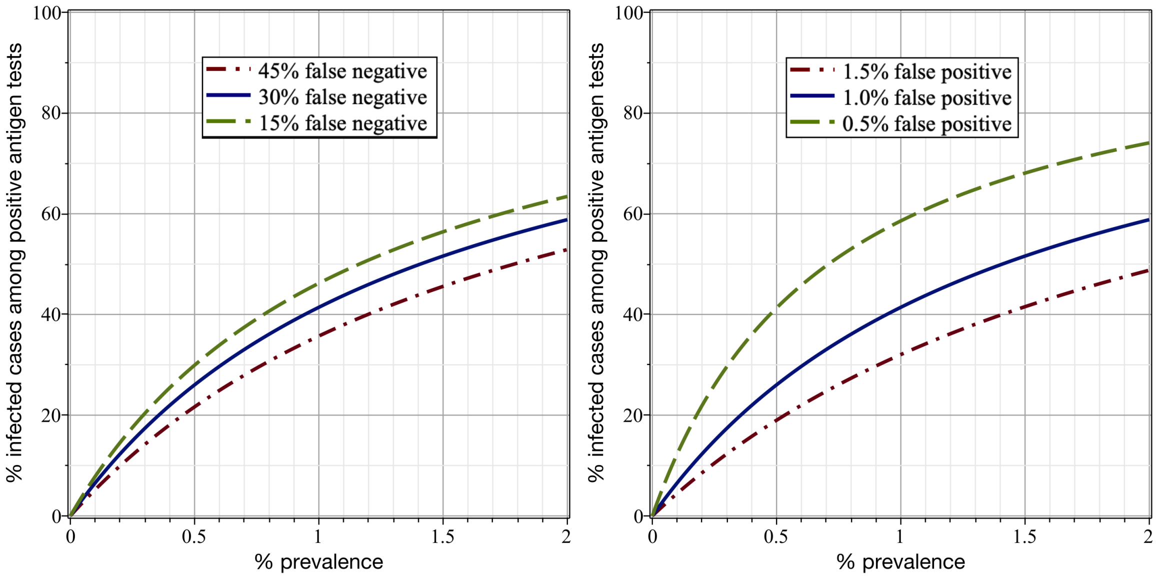


**Fig. S 6.** Fraction of infected cases among all people with positive virus antigen test result as function of prevalence. Left: for 15%, 30% and 45% false negative rates and a fixed false positive rate of 1%; right: for 0.5%, 1% and 1.5% false positive rates and a fixed false negative rate of 30%. The fraction of actually infected cases among positively tested individuals can be calculated as $p(1-f_n)/(p(1-f_n)+(1-p)f_p)$.

[8] Nathan W Furukawa, John T Brooks, and Jeremy Sobel. Evidence supporting transmission of severe acute respiratory syndrome coronavirus 2 while presymptomatic or asymptomatic. *Emerging infectious diseases*, 26(7), 2020.

[9] Hossein Gorji, Markus Arnoldini, David F Jenny, Wolf-Dietrich Hardt, and Patrick Jenny. Stecc: Smart testing with contact counting enhances covid-19 mitigation by bluetooth app based contact tracing. *medRxiv*, 2020.

[10] Shaobo He, Yuexi Peng, and Kehui Sun. Seir modeling of the covid-19 and its dynamics. *Nonlinear Dynamics*, pages 1–14, 2020.

[11] Ayaan Hossain, C. Alexander Reis, Sarthok Rahman, and M. Howard Salis. A massively parallel covid-19 diagnostic assay for simultaneous testing of 19200 patient samples, 2020.

[12] Can Hou, Jiaxin Chen, Yaqing Zhou, Lei Hua, Jinxia Yuan, Shu He, Yi Guo, Sheng Zhang, Qiaowei Jia, Chenhui Zhao, et al. The effectiveness of quarantine of wuhan city against the corona virus disease 2019 (covid-19): A well-mixed seir model analysis. *Journal of medical virology*, 2020.

[13] Enrico Lavezzo, Elisa Franchin, Constanze Ciavarella, Gina Cuomo-Dannenburg, Luisa Barzon, Claudia Del Vecchio, Lucia Rossi, Riccardo Manganelli, Arianna Loregian, Nicolò Navarin, et al. Suppression of covid-19 outbreak in the municipality of vo, italy. *medRxiv*, 2020.

[14] Ruiyun Li, Sen Pei, Bin Chen, Yimeng Song, Tao Zhang, Wan Yang, and Jeffrey Shaman. Substantial undocumented infection facilitates the rapid dissemination of novel coronavirus (sars-cov2). *Science*, 2020.

[15] Zhonghua Liu, Xing Bing, and Xue Za Zhi. The epidemiological characteristics of an outbreak of 2019 novel coronavirus diseases (covid-19) in china. *Novel Coronavirus Pneumonia Emergency Response Epidemiology Team*, 41(2):145, 2020.

[16] Ke Men, Xia Wang, Yihao Li, Guangwei Zhang, Jingjing Hu, Yanyan Gao, and Henry Han. Estimate the incubation period of coronavirus 2019 (covid-19). *medRxiv*, 2020.

[17] Anca Radulescu and Kieran Cavanagh. Management strategies in a seir model of covid 19 community spread. *arXiv preprint arXiv:2003.11150*, 2020.

[18] Dimple D Rajgor, Meng Har Lee, Sophia Archuleta, Natasha Bagdasarian, and Swee Chye Quek. The many estimates of the covid-19 case fatality rate. *The Lancet Infectious Diseases*, 20(7):776–777, 2020.

[19] Christina Savvides and Robert Siegel. Asymptomatic and presymptomatic transmission of sars-cov-2: A systematic review. *medRxiv*, 2020.

[20] Cormac Sheridan. Fast, portable tests come online to curb coronavirus pandemic. *Nat Biotechnol*, 10, 2020.

[21] Victor Virlogeux, Vicky J Fang, Joseph T Wu, Lai-Ming Ho, JS Malik Peiris, Gabriel M Leung, and Benjamin J Cowling. Incubation period duration and severity of clinical disease following severe acute respiratory syndrome coronavirus infection. *Epidemiology (Cambridge, Mass.)*, 26(5):666, 2015.

[22] Joseph T Wu, Kathy Leung, and Gabriel M Leung. Nowcasting and forecasting the potential domestic and international spread of the 2019-ncov outbreak originating in wuhan, china: a modelling study. *The Lancet*, 395 (10225):689–697, 2020.

[23] Fei Zhou, Ting Yu, Ronghui Du, Guohui Fan, Ying Liu, Zhibo Liu, Jie Xiang, Yeming Wang, Bin Song, Xiaoying Gu, et al. Clinical course and risk factors for mortality of adult inpatients with covid-19 in wuhan, china: a retrospective cohort study. *The Lancet*, 2020.
